# How nurses spend their time: nurses’ experiences and time use for providing HIV treatment under conventional and differentiated service delivery models in South Africa

**DOI:** 10.64898/2026.06.06.26355033

**Authors:** Nkgomeleng A. Lekodeba, Sophie J.S. Pascoe, Amy N. Huber, Nkosinathi Ngcobo, Allison J. Morgan, Vinolia Ntjikelane, Anushka R. Marri, Linda Sande, Khumbo Shumba, Idah Mokhele, Brooke E. Nichols, Lise Jamieson, Sydney Rosen

## Abstract

**Introduction:** Differentiated service delivery (DSD) models aim to reduce time healthcare providers spend with DSD clients, increasing time available for non-DSD clients. We measured nurses’ time allocation and explored their experiences with DSD models in South Africa.

**Methods:** We conducted time and motion observations and surveyed nurses at 24 public primary healthcare facilities across two SENTINEL study rounds (09/2022-07/2023 and 11/2023-07/2024). We report median time nurses spent by activity, model of care, and interaction type. Log binomial regression investigated factors associated with high direct nurse-client interaction (above median minutes) and extended work-days (≥9 hours), and estimated adjusted risk ratios (aRR). Survey questions were related to client care, additional time availability, and policy changes post DSD implementation, with key themes presented alongside illustrative quotes.

**Results:** 176 nurses (88% female, median age 44) were observed for 344 working days; of these, 60 (34%) participated in the provider survey. Nurses spent a median of 293 minutes (53% of their work-day) on direct nurse-client interaction, 89 minutes (22%) on client-support or facility-related tasks, and the remainder on other activities including personal breaks. Time spent per client was similar across conventional care clients (11 [IQR: 8-15] minutes) but ranged between 9 (7-13) to 11 (8-15) minutes for DSD clients; number of direct nurse-client interactions did not differ meaningfully. Nurses at facilities with 2,000-3,999 total remaining on ART (TROA) (aRR 1.56, 95% CI: 1.02-2.37) and in urban areas (aRR 1.43, [1.08-1.89]) had more direct nurse-client interactions than those at facilities with <1,999 TROA and in rural areas, respectively. Nurses at facilities with 4,000+ TROA (aRR 2.22, [1.36-3.63]) and those observed in SENTINEL 3.0 (aRR 1.53, [1.13-2.07]) were more likely to work standard or longer workdays than those at lower TROA facilities (<1,999), those in SENTINEL 2.0 and urban areas. Nurses reported DSD models improved client care (90%), freed up time (60%), and changed clinic procedures and policies (60%).

**Conclusions:** While DSD models did not significantly reduce direct nurse-client interaction time, nurses reported improved client care and gained additional time. DSD impact may vary by facility context. As DSD implementation expands, effective time reallocation may enhance facility performance and provider productivity.

## Introduction

The introduction and scale up of differentiated service delivery (DSD) models for HIV treatment in sub-Saharan African countries was expected to produce a wide range of positive changes for health systems, healthcare providers, and antiretroviral therapy (ART) clients [1]. Most DSD models require fewer clinic visits for “established” treatment clients, who have been on ART for at least 6 or 12 months and whose viral load is suppressed, than does conventional (non-differentiated) care. DSD models that reduce facility resource utilization per clients, often called “low intensity” models, include multi-month dispensing of medications, community-based pickup points for medication refills, and “fast track” procedures that allow patients to refill prescriptions at facilities without undergoing a full clinical visit.

As larger proportions of established ART patients transitioned to DSD models in recent years, anticipated benefits included the decongestion of primary healthcare clinics [2–4] and more provider time available for attending to non-established ART clients and other chronic disease clients and for conducting community outreach activities [5]. Guidelines generally do not recommend how healthcare providers should spend their time, however. Providers in Malawi, Zambia, and South Africa have self-reported a reduction in work burden [6-8] after DSD model implementation, but there have been few real-world observations of healthcare providers’ time use since DSD models became common. The extent to which DSD uptake affects providers’ time allocations and workloads remains unclear.

Given the well-documented problems with self-reporting one’s own time use [8,9], direct observation offers a more accurate method to evaluate how DSD models have altered human resource time allocation at healthcare facilities. As part of the SENTINEL study [10] in South Africa, we conducted time and motion observations to measure the impact of DSD implementation on nurses’ time use and administered surveys to explore their post-DSD perceptions of their time use in South African primary public health facilities, a sub-set of which is included in the analyses here [7].

## Methods

### Study design and sites

The SENTINEL study was conducted at 24 primary healthcare facilities in four South African districts: Alfred Nzo (Eastern Cape Province), Ehlanzeni (Mpumalanga Province), King Cetshwayo (KwaZulu-Natal Province), and West Rand (Gauteng Province). For this analysis we used data from two of three SENTINEL rounds: SENTINEL 2.0 from 13 September 2022 to 24 July 2023 and SENTINEL 3.0 from 29 November 2023 to 17 July 2024. SENTINEL 1.0 data were excluded due to substantial differences in data type and collection instruments compared to those in subsequent rounds, which would have compromised dataset comparability and reporting. SENTINEL study sites were purposively selected to provide sufficient ART client volumes for survey sample size, variation in setting (rural or urban), and mix of DSD models for HIV treatment. All SENTINEL facilities in South Africa employ a “single-stream” model of chronic disease care, whereby patients with chronic conditions, including HIV, are integrated within the same service pathway. Under this model, all chronic care clients share common queues and are attended to sequentially by the same healthcare providers, irrespective of their specific chronic condition. The SENTINEL study has been described in detail elsewhere [10], and details of facility characteristics are provided in Supplementary Table S1. (We note that South Africa refers to its DSD models as “Differentiated Models of Care” or DMOCs; we use DSD models here to align with terminology commonly used in other country contexts.)

Each study site offered two main DSD models at the time of our study: facility-based pick-up points and external pick-up points. Both were supplied by South Africa’s Central Chronic Medicines Dispensing and Distribution (CCMDD) program, which pre-packs medication parcels at central locations and delivers them to clients with chronic diseases either at facilities (facility pickup points) or at community locations such as private retail pharmacies (external pickup points) [11,12]. Some facilities also offered one or more other models, such as adherence clubs, electronic medication lockers, youth clubs, and home ART delivery. At the time of the study, eligibility requirements for DSD enrollment included at least six months’ experience on ART and documentation of a suppressed viral load.

All facilities continued to offer conventional (non-differentiated) care for clients not enrolled in a DSD model. We divided conventional care into two groups: 1) eligible for DSD but not enrolled, and 2) not eligible for DSD and not enrolled. As previously described [13], eligible clients must be registered for DSD models by providers, at their own request and/or the provider’s initiative. While enrolment of eligible clients is strongly encouraged by the National Department of Health, facilities differ widely in DSD uptake and enrollment. As of March 2024, approximately 49.5% of eligible clients in South Africa were reported to be enrolled in a DSD model, leaving the rest in conventional care [14].

### Participants and data collection

#### Study recruitment and enrolment

For this analysis, we used data from two components of the SENTINEL study: 1) time and motion observations of up to 5 healthcare providers for up to two working days per facility; and 2) surveys with up to 10 healthcare providers per facility. The analytic sample was restricted to nurses who participated in time and motion observations. Where a nurse also completed the survey, their responses were linked using a unique study identification number and included in the analysis.

Providers participating in the survey and for time and motion observations, were purposively selected by trained research assistants with the help of the facility operations manager. Providers were eligible if they had direct or indirect involvement with the facility’s implementation of ART and DSD models and had been employed at the facility for at least six months. Trained research assistants introduced the study and potential participants were then invited individually in a private area within the facility to learn more about the study processes and provide written informed consent. Demographic characteristics, professional cadre (rank), and role within the facility were documented for each participant.

#### Time and motion observations

Nurses were observed on weekdays representative of typical ART client care days. The nurse work-day was defined as a 9-hour (540-minute) standard period in accordance with South African labour laws [15]. A research assistant stationed outside the participant’s primary workspace (e.g. consultation room) recorded the start and end times of each activity or interaction throughout the full workday. Research assistants were trained to remain minimally intrusive and not to disrupt the regular flow of clinical activities. Activities were broadly categorized as clinical consultations, client support or facility related tasks, or other activities (Table 1).

**Table 1:** Time block uses included in analysis.

| Category | Activity sub-categories | Description |
| --- | --- | --- |
| Clinical consultations | Direct nurse-client interaction | General clinical consultation with clients or provider-client interaction for clinical procedures such as laboratory testing/sample collection or medication refills |
| Client-support or facility related tasks | Client-related | Tasks such as client waiting que delegation, record keeping or retrieving client information |
|  | General administration or meetings | Performing administrative activities or attending facility and staff-related meetings |
| Other activities | Personal breaks | Personal breaks such as lunch/tea breaks and other personal time such as taking personal calls |
|  | Involuntary free time | No direct provider-client interactions due to lack of clients and no assigned tasks or other activities |
|  | Not on site | Not present at the study facility during the prescribed work-day and not attending to any facility related task. |

To respect client privacy, research assistants did not did not enter consulting rooms during consultations or directly observe interactions between nurses and clients. Instead, participating nurses completed a brief checklist during each direct consultation with a client. The checklist captured client type (initiating ART, 0-6 months on ART, >6 months on ART, non-HIV), service conducted during the visit (medication refill, laboratory tests, HIV testing); and the DSD model in which the client was enrolled, if any. Research assistants retrieved and entered these checklists into the participant’s main study record after each consultation was completed (Supplementary file 1).

#### Participant survey

The SENTINEL provider survey included closed- and open-ended questions about providers’ experiences since the implementation of DSD models (Supplementary file 2). Participants were prompted to elaborate on their responses to gain deeper insights into how DSD implementation affected their workdays. For this analysis, we utilized SENTINEL 2.0 survey questions pertaining to workloads and time use, and SENTINEL 3.0 survey responses were excluded as they did not include these questions. A broader set of provider experiences of DSD implementation from the first round of SENTINEL are reported elsewhere [7,10].

### Data analysis and outcomes

Each study facility provided estimates of the number of ART clients it served (total remaining on ART, abbreviated TROA) and the number of clients enrolled in each DSD model at the time of SENTINEL rounds 2 and 3, based on electronic medical records (EMR) and facility-level DSD client registers. We calculated DSD uptake as the proportion of patients currently on ART at each facility, as indicated in the EMR, who were enrolled in DSD, based on the registers. To stratify facilities by size (total retained in care, TROA) and DSD uptake, we estimated the median for each variable across both rounds of SENTINEL data and classified each facility as above or below that median. Facilities falling below the median TROA or DSD uptake were classified accordingly, with the same nomenclature applied consistently across both variables. Facility-level data used to calculate these medians are provided in Supplementary Table S1.

Characteristics of participants were described using frequencies, proportions, and medians with interquartile ranges (IQR) as appropriate. For the time and motion analysis, we estimated the median (IQR) time (in minutes) spent by nurses per observed nurse-workday for each of their activities or interactions. We then analyzed the distribution of nurse time throughout the course of the workday, stratified by DSD uptake. We estimated the median number of client interactions and median minutes spent per client and per type of healthcare interaction, disaggregated by model of care.

We used binomial regression with a logarithmic link function to investigate factors associated with high direct nurse-client interaction and standard work hours, and reported crude and adjusted risk ratios (aRR) [16]. Direct nurse-client interaction was categorized using the sample median minutes: nurses with direct nurse-client interaction below the median were classified as having low interaction, and those with above median time as having high interaction. Work hours per day were categorized as <540 minutes (less than standard work hours) or ≥540 minutes (standard or more work hours). Models were adjusted using a stepwise regression approach for facility size-TROA, % pooled median DSD uptake, facility location, number of days per week providing HIV services, and the SENTINEL study period (round 2 vs 3).

For the provider survey analysis, we calculated the proportion of providers who reported changes in client care practices, availability of free or additional time, and changes to clinic procedures or policies due to the implementation of DSD models. Open-ended responses were coded in Excel by one study team member. We utilized an inductive approach to conduct a content analysis and identify emergent themes [17]. We then used the emergent themes to guide which variables to look at from qualitative data. Key themes were summarized and presented alongside illustrative quotes.

### Ethics review

The SENTINEL study was reviewed and approved by the Human Research Ethics Committee (Medical) of the University of Witwatersrand (South Africa), protocol number M210241; and the Boston University IRB (United States), protocol number H-41402. All participants provided written informed consent. SENTINEL is registered on clinicaltrials.gov (NCT05886530) and the South African National Clinical Trial Registry (DOH-27-052023-4669). National Health Research Database approval for conducting the survey was provided for each district.

## Results

### Participant characteristics

A total of 176 nurses participated in the time and motion observations and were included in the analysis from both rounds of SENTINEL, including 60 nurses who also completed the provider survey (Table 2). Most participants were female (88%), with a median age of 44 years, median experience of 11 years in their current roles, and median employment at the study site of 5 years. Slightly more than half (53%) were employed at urban facilities.

**Table 2.**
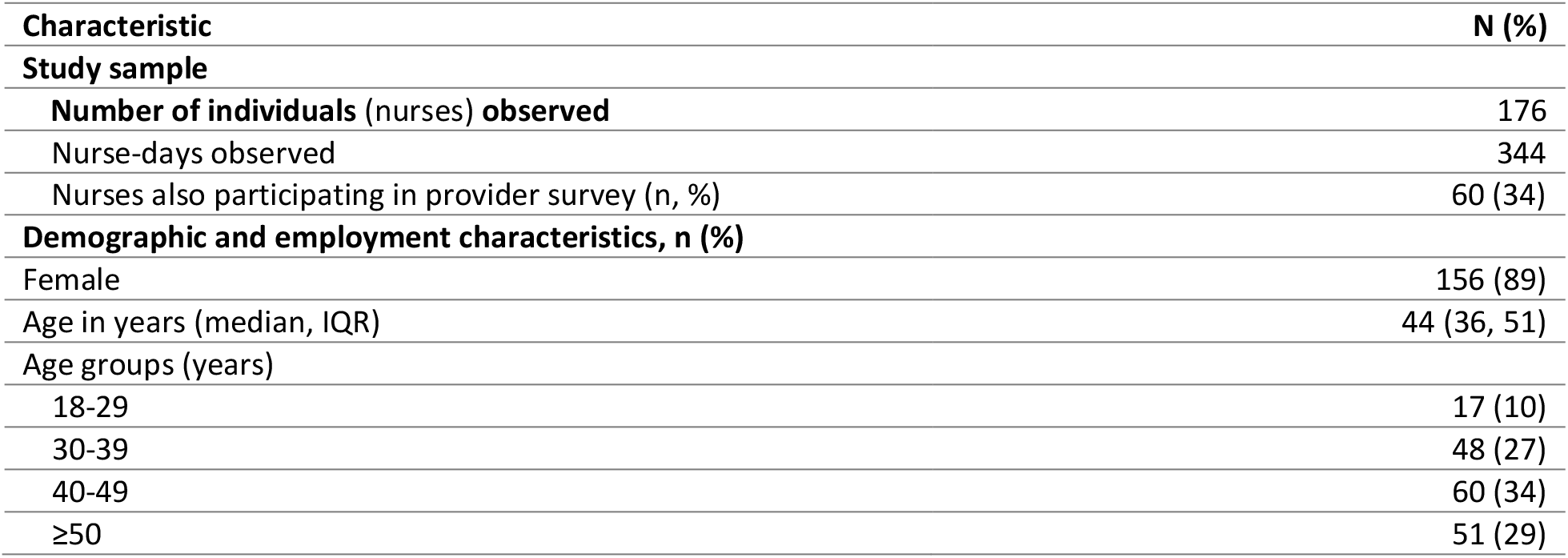
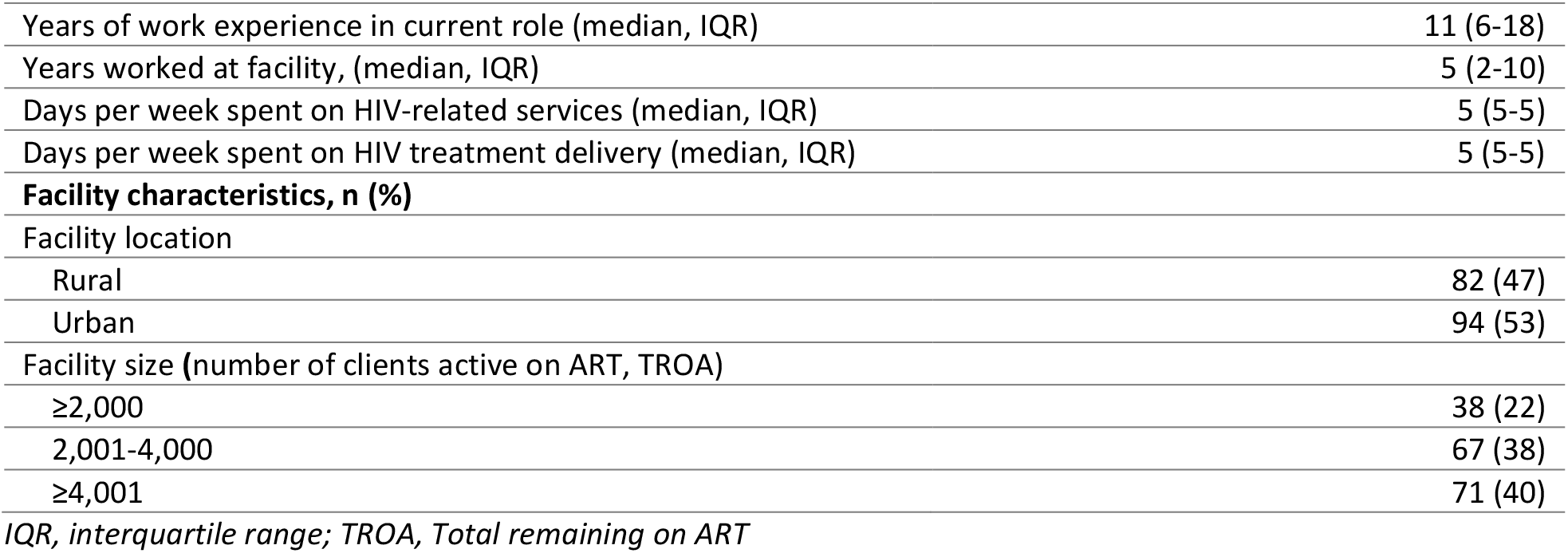
Participant and facility characteristics.

### Time allocation

Nurses spent a median of 293 minutes, just over half (53%) of the observed workday, on direct nurse-client interactions (Table 3, Fig 1). Client support tasks, which included client-related and DSD activities, took a median of less than 30 minutes per workday. Facility-related or other activities such as administrative tasks and meetings accounted for 60 minutes. Consistent with current labor practices in South Africa, nurses spent one hour on personal breaks. In addition, nurses had a median of 10 minutes of involuntary free time and 18 minutes of time not on site.

**Table 3.** Distribution of nurses’ time spent per interaction or activity for each work-day observed (pooled analysis)

| Characteristic | Number of activities or interactions | Daily time spent in minutes<sup>†</sup> (median, IQR) | % of daily time |
| --- | --- | --- | --- |
| **N (participants)** |  | 176 |  |
| **Days observed (n)** |  | 344 |  |
| **Interaction or activity** |  |  |  |
| Direct nurse-client interaction | 9,525 | 293 (237-345) | 53.2 |
| Client related tasks | 1,377 | 29 (7-72) | 9.1 |
| General administration or meetings | 776 | 60 (34-95) | 13.1 |
| Personal breaks | 530 | 64 (52-86) | 12.3 |
| Involuntary free time | 572 | 9 (0-50) | 6.0 |
| Not on site<sup>‡</sup> | 220 | 18 (0-45) | 6.1 |
| **Total work-day observed** | 13,000 | 540 (540-540) | 100 |

<sup>†</sup>The daily time spent in minutes on each interaction or activity does not add up to the total work-day (540 minutes) because these values represent medians, not cumulative totals.
<sup>‡</sup>Time when nurses were "not at site" was calculated by subtracting the duration of their actual observed work-day from the standard 540 minutes (9-hour) workday.

**Figure 1.**
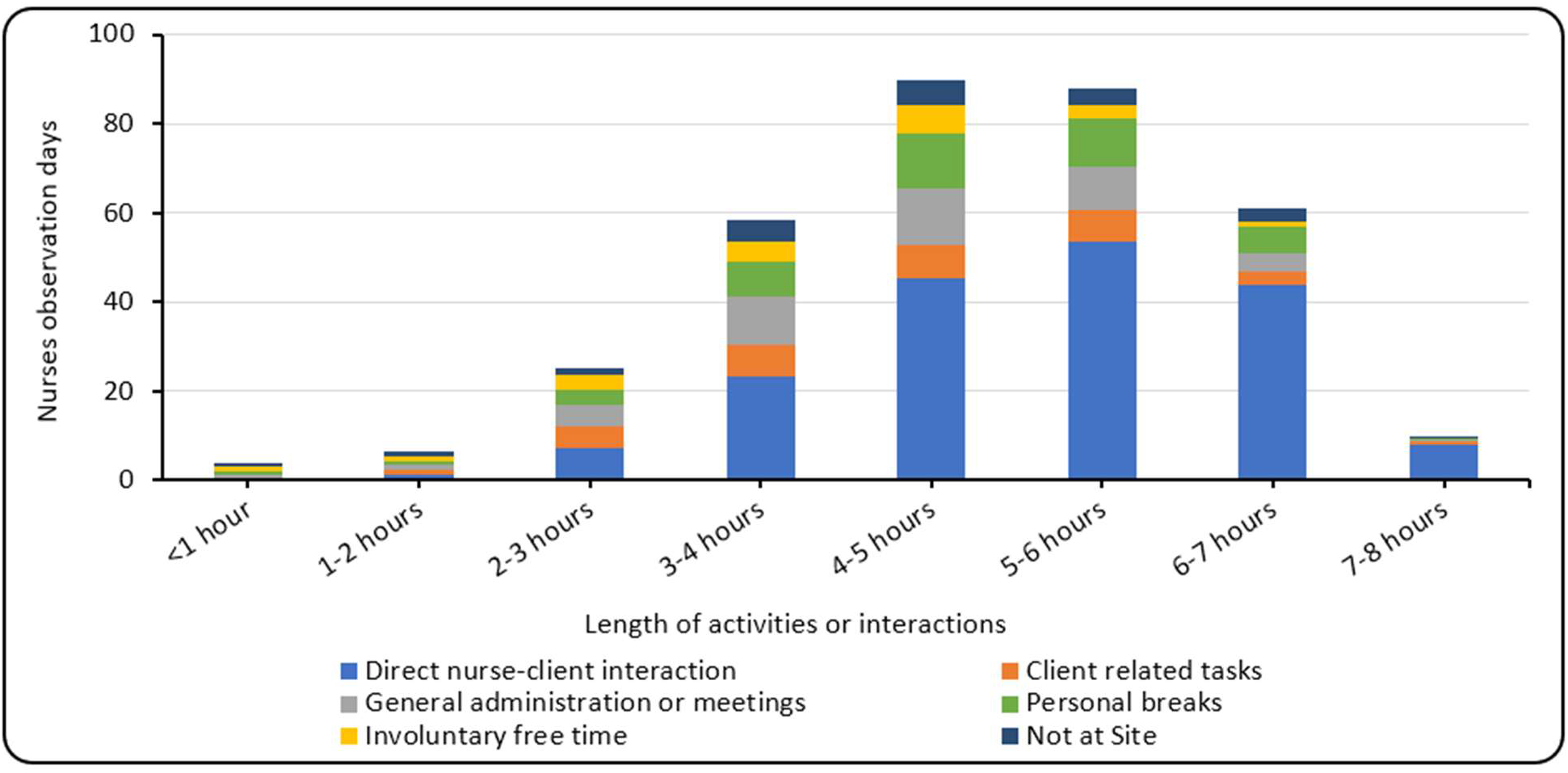
Nurses’ observed hours by the type of activity or interaction (histogram showing nurses observations days and the length of activities or interactions per workday)

### Time spent by model of care

Nurses had a median of 25 (IQR: 19-33) total client interactions per day, with a median of 11 (9-14) minutes per interaction. Roughly a fifth of direct nurse-client interactions were with clients in conventional care eligible for DSD and another fifth with clients in conventional care not eligible for DSD. There were slightly fewer interactions per day with clients enrolled in facility and external pickup points. The nurses in our study also interacted frequently (a median of 8 times per day) with non-HIV clients, reflecting South Africa’s single stream approach to chronic disease care. Approximately 33% of nurse time each day was spent on interactions with clients in DSD models, 43% on clients in conventional care, and 24% with other types of clients (Table 4).

**Table 4.**
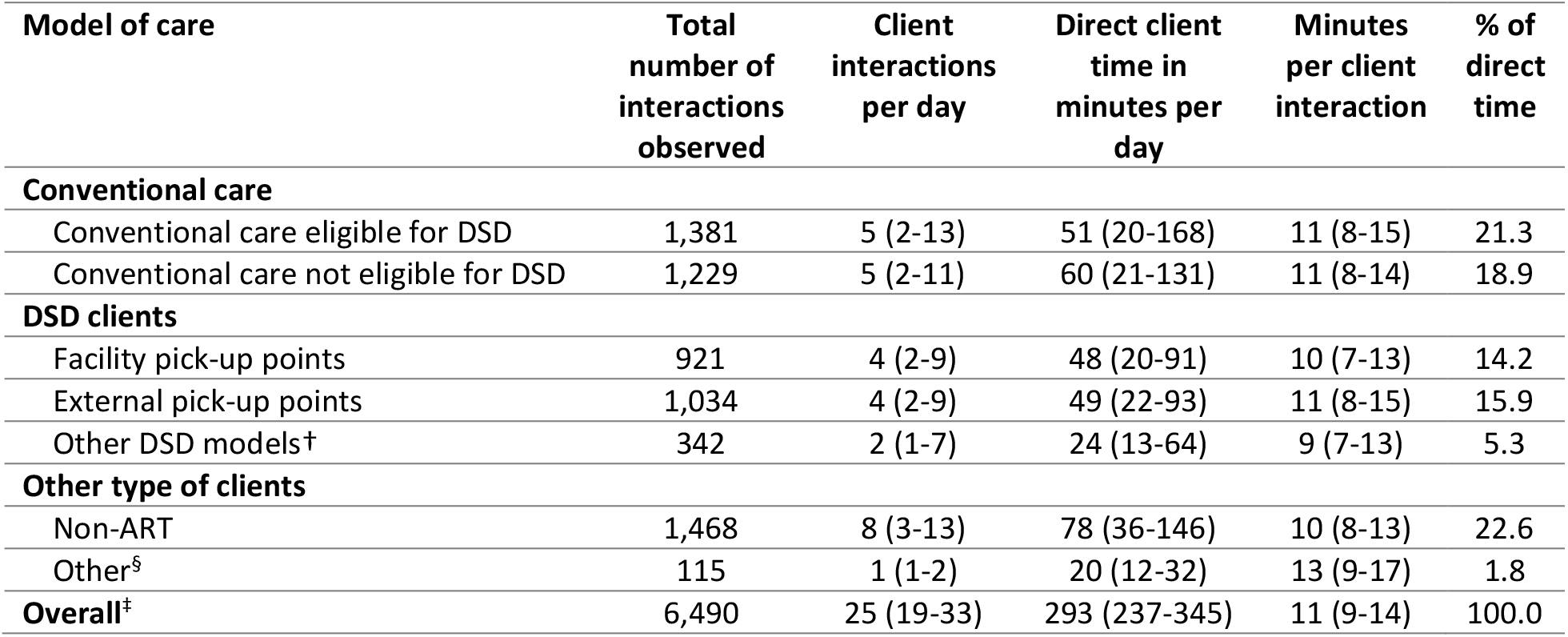
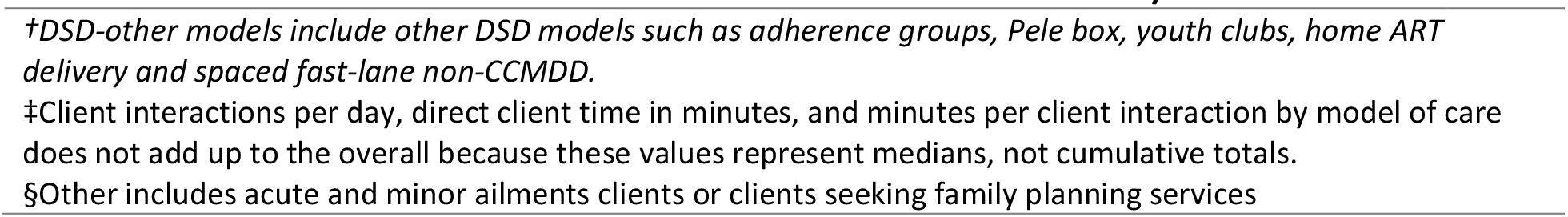
Distribution of direct nurse-client interactions by the type of model of care per nurse-workday observed (n=344 days) (median, IQR)

### Time spent by type of healthcare interaction

Nurses spent a median of 11 minutes per full clinic consultation for clients in conventional care and eligible for DSD (Fig 2, Table S2). As might be expected, they spent considerably longer –a median of 16 minutes –per full clinic consultation for clients in conventional care not eligible for DSD. Perhaps unexpectedly, consultations with clients enrolled in DSD models were longer than for those not enrolled, at a median of 19 minutes for those in facility pickup points and 17 minutes for those in external pickup points. Full clinic consultations were longest among non-ART clients and other client types, lasting 27 to 31 minutes per client. Laboratory test and medication collection interaction durations were comparable across conventional care, facility, and external pick-up point clients. Medication collection and re-scripting interactions occurred exclusively among DSD clients and lasted 9 to 11 minutes per client.

**Figure 2.**
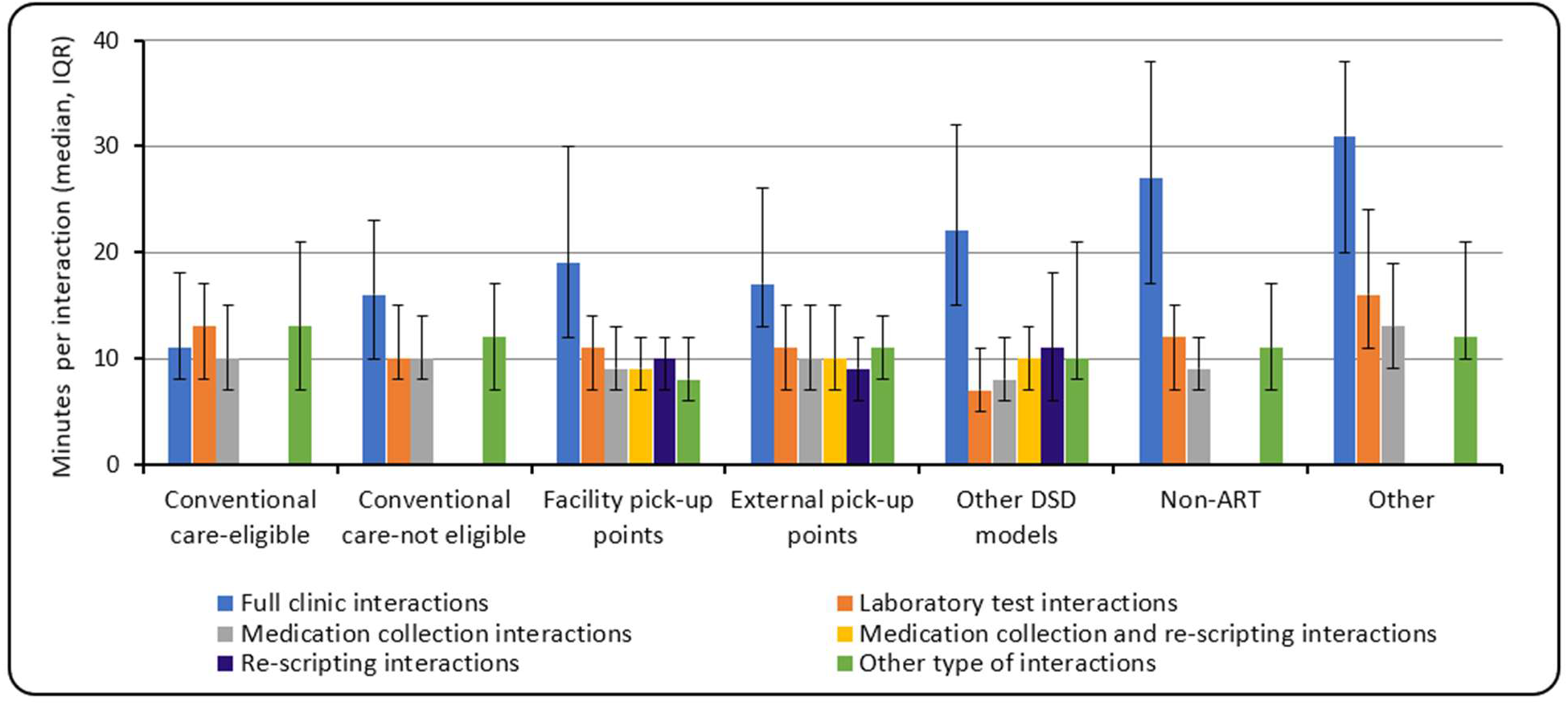
Minutes spent per model care and the type of healthcare interaction per nurse-day observed (median, IQR)* *\*Error bars on the figure represent interquartile ranges (IQR)*

### Factors associated with high numbers of direct nurse-client interactions and worked hours

Crude and adjusted relative risks of factors associated with high numbers of (a) direct nurse-client interactions per day, and (b) nine-or-more hour workday. Nurses at facilities with 2,000-3,999 TROA (aRR 1.56 [95% CI: 1.02-2.37] vs 4,000+ TROA aRR 1.24 [0.78-1.97]) and those in urban areas (aRR 1.43 [1.08-1.89]) were more likely to have higher numbers of direct nurse-client interactions per day than those with TROA of <1,999 and in rural areas, respectively (Fig 3a). There were no other factors that were associated with high or lower direct nurse-client interaction per nurse work-day observed (Fig 3a).

**Figure 3:**
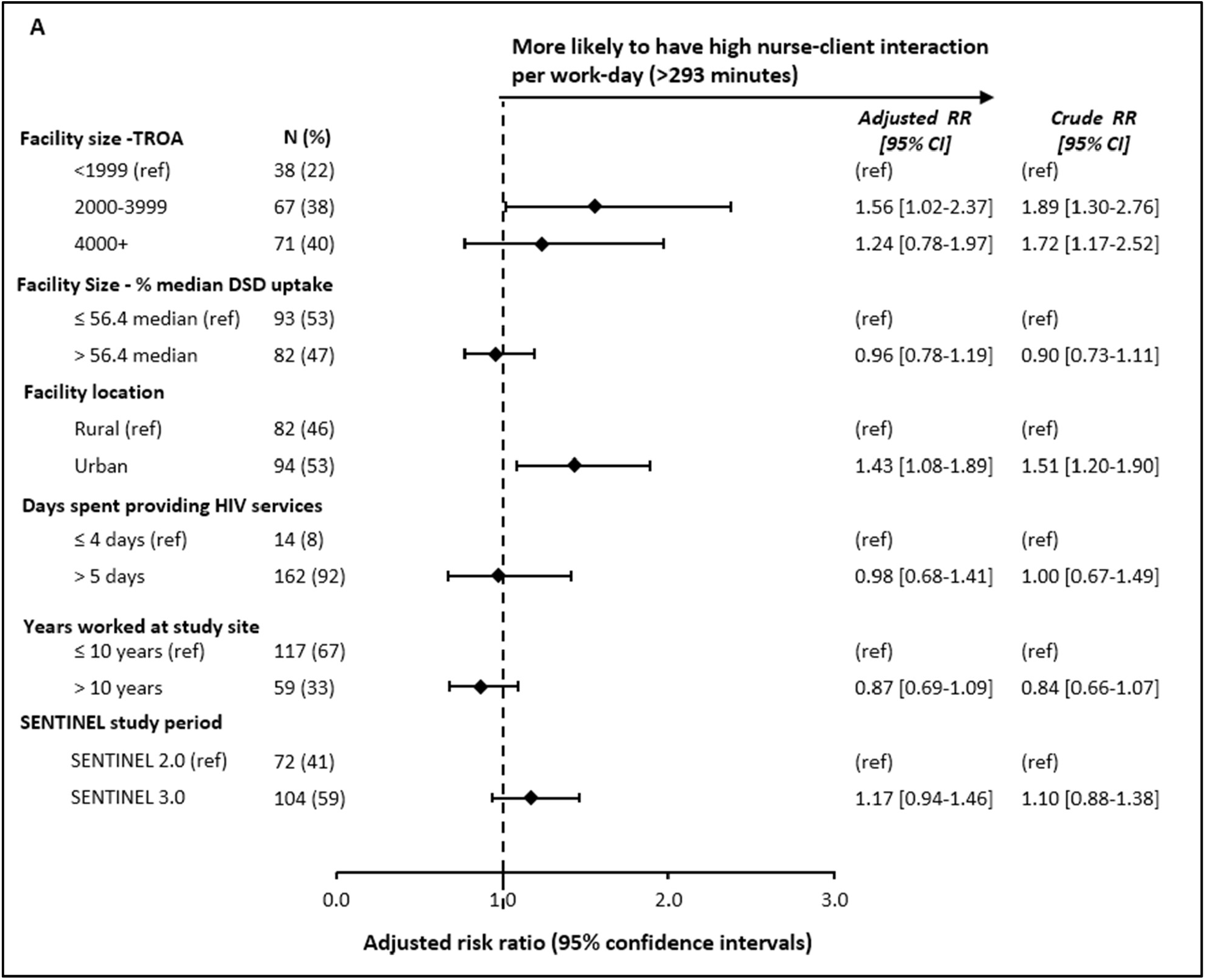
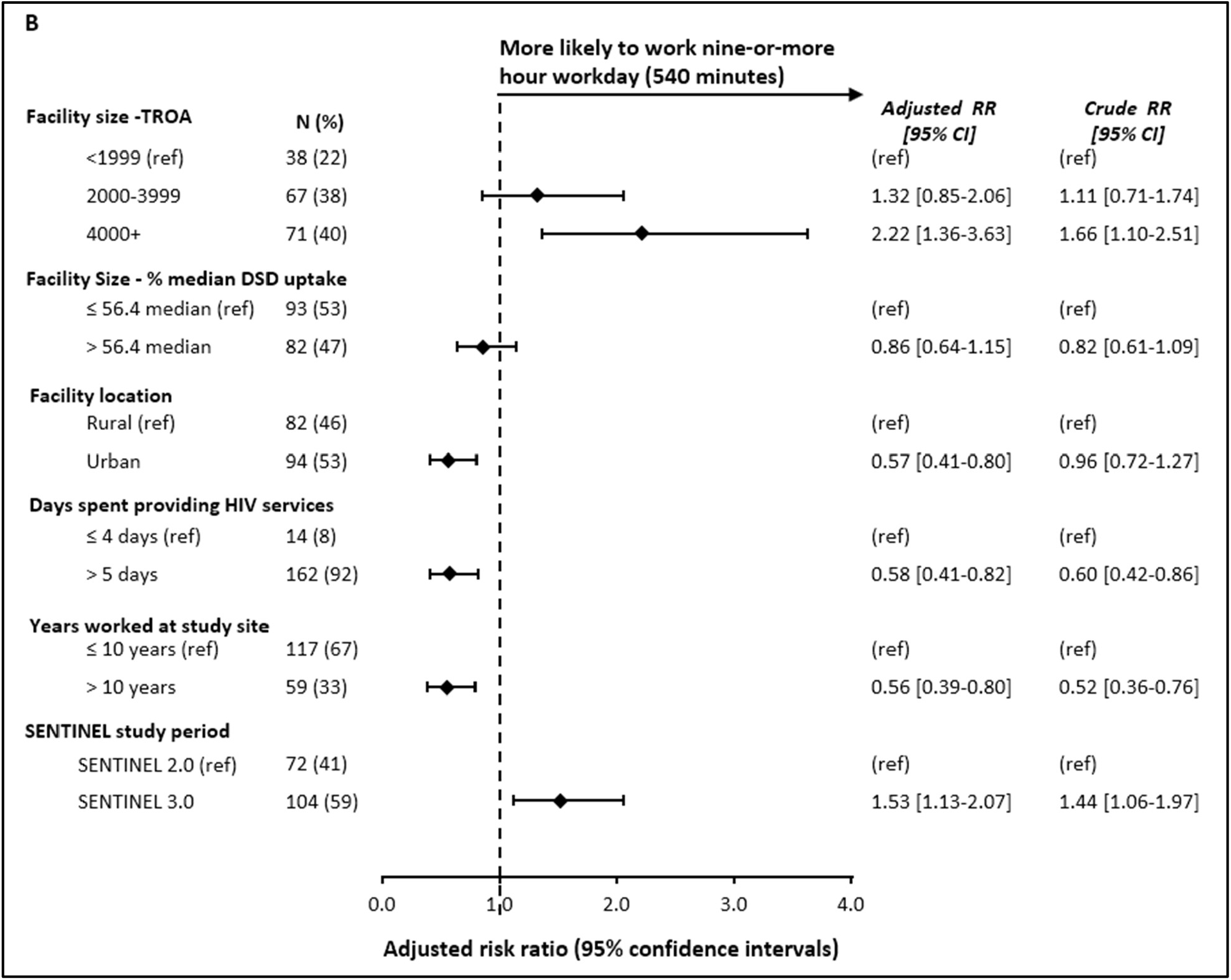
Crude and adjusted risk ratios for A) high number of direct nurse-client interactions per day (>293 minutes per nurse per day) and B) nine-or-more hour workday; adjusting for facility size - TROA, and % median DSD uptake, facility location, days spend providing HIV services in a week, years worked at facility and SENTINEL study period *TROA, Total remaining on ART; DSD: differentiated service delivery

Nurses in facilities with TROA of 4,000+ (aRR 2.22 [1.36-3.63] vs 2,000-3,999 TROA aRR 1.32 [0.85-2.06]) and those observed in SENTINEL round 2 (aRR 1.53 [1.13-2.07]) were more likely to have worked a standard workday than did those in facilities with lower TROA (<1,999) and those observed in SENTINEL round 1, respectively (Fig 3b). Working hours did not significantly differ for those staff at facilities with higher or lower DSD uptake or at facilities with less than 4,000 clients on ART. Nurses who worked in urban areas (aRR 0.57 [0.41: 0.80]), who supported provision of HIV services every day (aRR 0.58 [0.41-0.82]), or who were more experienced (aRR [0.39-0.80]) were less likely to work a standard work-day (Fig 3b).

### Provider experience

In the quantitative survey, most participants (60%) reported having additional free time as a result of DSD, while the rest reported not having additional free time (38%) or did not know (2%). Responses to qualitative, open-ended questions highlighted themes such as reduced workload, decongested clinics, and improved workflow efficiency (Table 5). Among providers who reported experiencing free or extra time, many appreciated that enrolling clients in DSD models allowed them to focus on other duties such as filing, completing paperwork, spending more time with clients, tracing clients and assisting in other departments. They also highlighted the ability to do other tasks such as ordering and pre-packing medication, conducting follow-ups with clients with unsuppressed viral loads and being able to prepare for the next day’s work in advance.

**Table 5.**
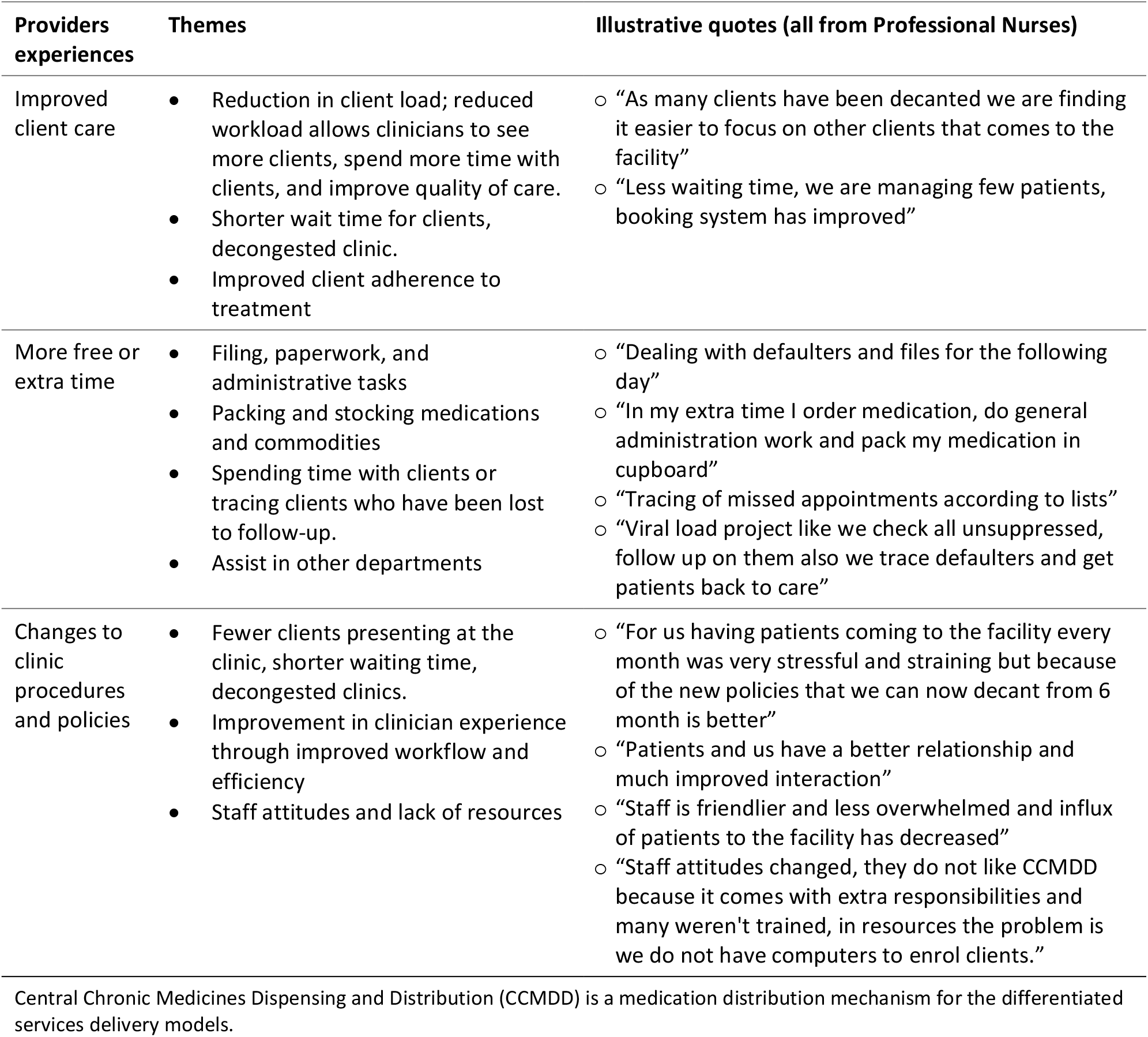
Provider views on time use after scaleup of differentiated models of care.

## Discussion

Low-intensity differentiated service delivery models for HIV treatment are expected to increase the time available for clinical care providers, primarily nurses, to spend with clients not enrolled in DSD models or requiring more complex care. In this study of nurses’ time use, we found little difference in the time nurses spent per client interaction between conventional care and the most common DSD models; the median interaction duration was 10-11 minutes, regardless of model of care. Time per interaction also does not appear to be influenced by the level of DSD uptake in a facility. Therefore, if DSD models are to meaningfully affect nurses’ time allocation, they must do so by changing the number of visits per year, rather than by changing how time is used during each visit.

The findings from the time and motion component of the study contrast fairly sharply with nurses’ own perceptions of the impact of DSD models on time use revealed in the survey, where 60% of respondents reported having time freed up since DSD implementation. We speculate that this may reflect the influence of outliers on participants’ perceptions of time use. Although overall time use and median interaction duration were similar regardless of DSD uptake, interquartile ranges were large, suggesting wide variation across individual providers, days, and direct client interactions.

To our knowledge, this is the first study to measure how nurses spend their time since the advent of large-scale DSD models in South Africa. Other studies have evaluated clients’ waiting time and compared differentiated services to conventional services [18,19]. A pre-DSD HIV-related time and motion study conducted in South Africa found that nurses spent a much larger majority of their time on direct nurse-client interaction (ranging from 82% to 84%) than we saw, while time per HIV client visit in that study ranged from 10 to 14 minutes, very similar to our results [20]. In contrast, a time and motion study in Zambia that was conducted during the scaleup of DSD models found that nurses spent less than half (46%) of their time on direct nurse-client interaction [21]. In Lesotho, average consultation durations were estimated at 19, 18, and 23 minutes for 3-month facility-based dispensing, 3-month dispensing at community adherence groups, and 6-month dispensing at community pick-up-points, respectively [22]. Similar to our qualitative findings, reduced workload and improved quality of care as a result of DSD implementation were self-reported by providers in Mozambique [23] and Zambia [24].

If, as we found in this study, the duration of nurse-client interactions does not differ consistently between low- and high-DSD uptake facilities, as well as between those enrolled in DSD models and conventional care, then the best opportunity for differentiated models of care to improve the efficiency of providers’ time allocations is by reducing the number of nurse-client interactions required per year. While we did not measure it in this study, the guidelines for South Africa’s facility and external pickup points allow clients enrolled in these models to have just two clinical consultations per year, rather than the four or more required for conventional models of care. Discussion of an even less intensive monitoring regime, involving 12-month scripting and one full clinical consultation per year, which South Africa permitted during the COVID-19 pandemic [25], is also underway. If adopted as an option and chosen by a substantial number of ART clients, this latter model has the potential to reduce nurse time used for maintaining patients on ART even more over a year.

The observation that nurses spend only just over half their time (54%) on direct nurse-client interaction also suggests room for improvement. Although South Africa has guidelines on the management of client waiting times across levels of care, including clinics and hospitals [26], to our knowledge, no policies exist establishing accepted norms for the number of client interactions per nurse per day or the proportion of time nurses should spend on direct nurse-client interaction versus other activities such as administrative duties and meetings [27,28]. This leaves nurses’ working patterns and time use unregulated and determined largely by clinic managers, who presumably vary in their performance of this task. Our finding underscores the need for policy action to optimize time distribution and reduce unproductive time.

Though our study provides valuable insights into nurses’ time use and their experiences of DSD implementation, our findings are subject to study design limitations that should be considered when interpreting the results. First, the use of median DSD uptake at the facility level to compare sites with “low” and “high” uptake may have missed more important thresholds of DSD participation at which nurses’ time use changes. It is certainly possible that DSD uptake exceeding 56% median among our study sites is needed to generate a measurable impact on time use. Second, and related to the first limitation, we lacked a pre-DSD comparison for our results. We cannot report the true pre/post impact of DSD implementation, only trends associated with greater or lesser uptake, which may in turn reflect other, unobserved facility characteristics. Third, the study was limited to a small number of study sites and participants in a single country, making generalizability across all of South Africa and to other countries uncertain. Fourth, we did not attempt to capture data on clients’ needs during consultations or participants’ activities during non-consultation time to assess the efficiency of time use, relative to demands on the participating clinics. And finally, the study took place before the recent global funding reductions that reduced donor-funded staffing at many facilities. Preliminary reports suggest that task-shifting is one of the main strategies used by facilities to make up for lost personnel, suggesting that current time use patterns may be somewhat different from those we observed in 2022 and 2023 [29,30].

We also note limitations associated with study data collection itself. A Hawthorne effect, in which study participants may have altered their normal daily duties due to observation, is possible [9]. Even though efforts were made to ensure privacy during direct nurse-client interactions by timing nurse-client interactions from outside the consultation rooms, the presence of research assistants may have influenced nurses to alter their behavior when with clients [9].

## Conclusions

In South Africa, higher and lower uptake of DSD models for HIV treatment does not appear to have a major effect on nurses’ time use per client or on the number of client interactions per day. Our study thus does not provide evidence to support the hypothesis that having more ART clients enrolled in DSD models will substantively alter nurses’ time use. That said, a majority of providers reported benefits such as decongested facilities, greater availability of free or additional time to do other activities, and changes in clinic procedures and policies. It also documents wide variation in time use, suggesting that efficiency may depend on facility characteristics, including potentially DSD uptake, and thus be amenable to improvement. Additional research will be needed to confirm the extent to which such self-perceived benefits have improved human resource allocation in the era of differentiated service delivery.

## Supplementary files

Supplementary table S1. Characteristics of the SENTINEL study sites

Supplementary file 1 AMBIT South Africa SENTINEL time and motion data collection tool

Supplementary file 2 AMBIT South Africa SENTINEL provider survey instrument

Supplementary table S2. Minutes by the type of healthcare interaction (median, IQR)

## Authors’ contributions

NAL, SR, AH and SP conceptualized the study. NAL, SR, AH and SP contributed to instrument design. NAL analysed the data and wrote the first draft. AJM coded and analyzed open ended responses. All authors reviewed and approved the final manuscript.

## Acknowledgements

We would like to thank the staff of the study sites for participating in the study South Africa National Department of Health for approving this research. We are particularly appreciative of the support of South African ministry of health DSD task team. We would also like to acknowledge other members of the AMBIT SENTINEL study team for their contributions to the project at large: Barbara Xhosa, Oratile Mokgethi, Nyasha Mutanda, Elizabeth Kachingwe, Aniset Kamanga, Prudence Haimbe, Timothy Tchereni, Andrews Gunda, Nyasha Mutanda, Frehiwot Birhanu, Hilda Shakwelele; Taurai Makwalu.

## Data availability statement

All data used in this study were collected by the study team following written informed consent. Data will be made publicly available in a data repository within one year of study closure, as approved by the supervising ethics committees.

