## Supplementary information Table S1 and Table S2 for "How nurses spend their time: nurses’ experiences and time use for providing HIV treatment under conventional and differentiated service delivery models in South Africa"

### *Site characteristics*

### Supplementary table S1. Characteristics of the SENTINEL study sites

| **Site** | **Facility location** | **Total remaining on ART**  **(TROA)** | | **% of ART patients enrolled in DSD models (DSD uptake)** | |
| --- | --- | --- | --- | --- | --- |
|  |  | **SENTINEL 2.0** | **SENTINEL**  **3.0** | **SENTINEL**  **2.0** | **SENTINEL 3.0** |
| ***Alfred Nzo*†** |  |  |  |  |  |
| Community Health Centre | Urban | - | 3,483 | - | 67.6% |
| Clinic | Rural | - | 1,394 | - | 62.4% |
| Clinic | Rural | - | 1,738 | - | 81.5% |
| Clinic | Urban | - | 4,103 | - | 83.4% |
| Clinic | Rural | - | 1,188 | - | 72.5% |
| Clinic | Urban | - | 7,014 | - | 56.9% |
| ***Ehlanzeni District*** |  |  |  |  |  |
| Clinic | Urban | 6,535 | 6,643 | 45.8% | 48.2% |
| Clinic | Rural | 3,773 | 3,618 | 60.1% | 70.6% |
| Clinic | Rural | 1,943 | 1,979 | 74.0% | 78.6% |
| Clinic | Rural | 3,002 | 3,097 | 77.8% | 79.1% |
| Community Health Centre | Urban | 6,224 | 6,848 | 38.1% | 35.4% |
| Community Health Centre | Urban | 5,517 | 5,778 | 44.8% | 39.8% |
| **West Rand district‡** |  |  |  |  |  |
| Clinic | Urban | 2,094 | 2,099 | 60.4% | 54.3% |
| Clinic | Rural | 2,261 | 2,509 | 40.5% | 35.5% |
| Clinic | Urban | 2,071 | 2,343 | 26.2% | 59.2% |
| Clinic | Urban | 2,843 | 2,829 | 53.1% | 61.4% |
| Clinic | Rural | 1,134 | 1,270 | 44.4% | 68.4% |
| Community Health Centre | Urban | 3,165 | 3,663 | 54.1% | 53.9% |
| ***King Cetshwayo District*** |  |  |  |  |  |
| Clinic | Rural | 1,312 | 1,432 | 72.1% | 55.9% |
| Clinic | Rural | 1,368 | 1,696 | 50.6% | 61.3% |
| Clinic | Rural | 2,584 | 2,648 | 47.5% | 40.4% |
| Clinic | Rural | 3,402 | 3,375 | 49.9% | 64.9% |
| Clinic | Rural | 5,531 | 5,168 | 46.6% | 50.1% |
| Clinic | Urban | 8,112 | 8,132 | 71.1% | 80.3% |
| †Facilities added in study period 2  ‡SENTINEL clinics in the West Rand District did not have non-governmental partners supported by PEPFAR. Sentinel clinics in the other districts received PEPFAR support from the Aurum Institute, Broadreach Healthcare, or Right to Care…etc. | | | | | |

### Supplementary Table S2. Minutes by the type of healthcare interaction (median, IQR)

| Type of healthcare interaction | Conventional care clients | | DSD clients | | | Other client type | |
| --- | --- | --- | --- | --- | --- | --- | --- |
|  | **Conventional care eligible for DSD** | **Conventional care not eligible for DSD** | **Facility pick-up points** | **External pick-up points** | **Other DSD models** | **Non-ART** | **Other** |
| Full clinic visit | 11 (8, 18) | 16 (10, 23) | 19 (12, 30) | 17 (11, 26) | 22 (15, 32) | 27 (17, 38) | 31 (20, 38) |
| Laboratory test | 13 (8, 17) | 10 (8, 15) | 11 (7, 14) | 11 (7, 15) | 7 (5, 11) | 12 (7, 15) | 16 (11, 24) |
| Medication collection | 10 (7, 15) | 10 (8, 14) | 9 (7, 13) | 10 (7, 15) | 8 (6, 12) | 9 (7, 12) | 13 (9, 19) |
| Medication collection and rescripting | 0 (0, 0) | 0 (0, 0) | 9 (7, 12) | 10 (7, 15) | 10 (7, 13) | 0 (0, 0) | 0 (0, 0) |
| Rescripting | 0 (0, 0) | 0 (0, 0) | 10 (7, 12) | 9 (6, 12) | 11 (6, 18) | 0 (0, 0) | 0 (0, 0) |
| Other type | 13 (7, 21) | 12 (7, 17) | 8 (6, 12) | 11 (8, 14) | 10 (8, 21) | 11 (7, 17) | 12 (10, 21) |
